# Universal PCR and antibody testing demonstrate little to no transmission of SARS-CoV-2 in a rural community

**DOI:** 10.1101/2020.08.15.20175786

**Authors:** Ayesha Appa, Saki Takahashi, Isabel Rodriguez-Barraquer, Gabriel Chamie, Aenor Sawyer, CLIAHUB Consortium, Elias Duarte, Jill Hakim, Keirstinne Turcios, Joanna Vinden, Owen Janson, Aashish Manglik, Michael J. Peluso, Steven G. Deeks, Timothy J. Henrich, Leonel Torres, Mary Rodgers, John Hackett, Charles Chiu, Diane Havlir, Bryan Greenhouse

## Abstract

**Background:** The absence of systematic surveillance for SARS-CoV-2 has curtailed accurate appraisal of transmission intensity. Our objective was to perform case detection of an entire rural community to quantify SARS-CoV-2 transmission using PCR and antibody testing.

**Methods:** We conducted a cross-sectional survey of the prevalence and cumulative incidence of SARSCoV-2 infection in the rural town of Bolinas, California (population 1,620), four weeks following shelter-in-place orders. Residents and county essential workers were tested between April 20^th^ – 24^th^, 2020. Prevalence by PCR and seroprevalence combining data from two forms of antibody testing were performed in parallel (Abbott ARCHITECT IgG to nucleocapsid protein and in-house IgG ELISA to the receptor binding domain).

**Results:** Of 1,891 participants, 1,312 were confirmed Bolinas residents (>80% community ascertainment). Zero participants were PCR positive. Assuming 80% sensitivity, it would have been unlikely to observe these results (p< 0.05) if there were > 3 active infections in the community. Based on antibody results, estimated prevalence of prior infection was 0.16% (95% CrI: 0.02%, 0.46%). Seroprevalence estimates using only one of the two tests would have been higher, with greater uncertainty. The positive predictive value (PPV) of a positive result on both tests was 99.11% (95% CrI: 95.75%, 99.94%), compared to PPV 44.19%-63.32% (95% CrI range 3.25%-98.64%) if only one test was utilized.

**Conclusions:** Four weeks following shelter-in-place, active and prior SARS-CoV-2 infection in a rural Northern California community was extremely rare. In this low prevalence setting, use of two antibody tests increased the PPV and precision of seroprevalence estimates.

## INTRODUCTION

During early months of 2020, the SARS-CoV-2 virus proliferated in various metro areas across the United States leading to devastating numbers of COVID-19 cases; unfortunately testing availability lagged behind disease spread.[1] Months into the nation’s epidemic, with numerous reports of asymptomatic and presymptomatic transmission published[2–5], systematic surveillance for SARS-CoV-2 was still absent, eliminating the possibility of containment and limiting mitigation efforts.[6] Widespread testing regardless of symptoms has been proposed as a way to better understand the epidemiology of disease and curtail transmission. For example, in Vo, Italy, repeated testing of an entire town identified a high prevalence of asymptomatic infections and facilitated a reduction in diagnosis of new cases by approximately 50%.[7]

In the US, testing primarily symptomatic patients has revealed stark demographic, clinical, and regional differences in number and severity of COVID-19 cases. Older age, male sex, and cardiovascular comorbidities are risk factors associated with COVID-19 requiring hospitalization, and Black and Latinx individuals have faced disproportionately higher rates of infection and death.[8,9] Regionally, there are also clear differences in the proportion of positive PCR tests, from 33% in New York state to 5% in Los Angeles, though varied testing criteria and limited testing availability, particularly in non-urban areas, make these data difficult to interpret.[10,11] Measuring antibodies to SARS-CoV-2 can capture infections missed by PCR, but many seroprevalence studies published to date have been constrained by accuracy concerns, particularly those relying on non-laboratory-based, point-of-care antibody testing.[12,13] Furthermore, the penetrance of COVID-19 into rural communities, which may be particularly vulnerable given a high proportion of elderly residents, is unknown.

In the first effort of its kind to offer testing for active and prior COVID-19 infection to an entire town, we sought to 1) perform active case detection in the general population to identify and isolate potential reservoirs of infection, and 2) estimate SARS-CoV-2 prevalence and seroprevalence in a Northern California rural community, using PCR and laboratory-based antibody testing to capture those with active and past infection. We hypothesized that prevalence would be relatively low, given relatively fewer reported cases in the Bay Area compared to other regions, and thus used two orthogonal antibody tests to ensure adequate specificity for assessing prior infection.

## METHODS

Four weeks following shelter-in-place health orders, we conducted active population-based surveillance for SARS-CoV-2 infection in Bolinas, California among residents over age 4 years and Marin county first responders and essential workers. In close partnership with the community and Marin Department of Public Health, oropharyngeal and mid-turbinate swabs for reverse transcription polymerase chain reaction (RT-PCR) testing and blood for antibody testing were collected over five days (April 20^th^ – April 24^th^, 2020) as previously described in detail and summarized below.[14]

### Study Setting & Community Partnership

Located less than 30 miles from the San Francisco metro area, Bolinas is a rural town 5.8 square miles in size, bounded by the Pacific ocean, a wilderness area, and a lagoon. The 2010 census estimated population was 1,620 persons with population density of 278 persons per square mile[15], while the American community survey (ACS) in 2018 estimated the population size to have declined to 1,077 persons, 46% percent of whom were aged 65 years and older.[16] The majority of the community is White/Caucasian (88%), including 2% Latinx, with 3% Asian/Pacific Islander and 9% reporting multiple races. The median annual household income was $57,708, and 17% of the community lived in poverty.[16]

This study was community-initiated and co-led by Bolinas community leaders, who contributed throughout the planning and operational process. Additionally, key community stakeholders, including the main community-based health organization (Coastal Health Alliance) and the Bolinas Fire Department, provided endorsement and operational support. Together with study leadership, these community leaders and stakeholders participated in a virtual Town Hall the week prior to testing and week following results provision to introduce the study to the community, provide education, answer questions, and address concerns.[14]

### Testing Procedures

We conducted four days of drive-through and walk-up testing. On the fifth day of testing, we conducted limited in-home testing for home-bound participants. To pre-register, participants completed an online consent and survey (available in both English and Spanish), which included questions related to demographics, movement information, and past and current symptoms. Alternatively, participants were able to register by phone or on-site, though survey data collection was limited for the latter category. The testing was performed at a centrally located outdoor location in town, and participants were encouraged to drive if able. Participants remained in their vehicle (or physically distanced from other participants if on foot) while medical staff first collected blood (300–500 microliters) for subsequent antibody testing using fingerstick collection, then performed oropharyngeal and mid-turbinate specimen collection for RT-PCR with spun polyester swabs, following recommended procedures from the UCSF clinical laboratory.

### Laboratory Assays

At a CLIA-certified laboratory operated by UCSF and the Chan Zuckerberg Biohub, RTPCR of SARS-CoV-2 N and E genes as well as human RNAse P gene was completed using a Laboratory Developed Test with a limit of detection of log_10_ 4.5 viral genome copies/mL. RT-PCR specimens were stored in DNA/RNA Shield (Zymo Research) to inactivate virus and preserve RNA stability. Serum was obtained from fingerprick samples via centrifugation and stored at –20C until testing. Samples were tested using two independent assays: 1) the ARCHITECT SARSCoV-2 IgG immunoassay, for antibodies to the SARS-CoV-2 nucleoprotein (Abbott Laboratories, Abbott Park, IL, USA)[17], and 2) an in-house ELISA assay detecting IgG to the receptor binding domain of spike protein, based on published protocols.[18,19] For the ELISA, all samples which had an optical density (OD) above the cutoff (plate-specific OD for the CR3022 monoclonal antibody at 8 ng/uL), were repeated with titering to obtain an estimate of antibody concentration.

### Outcomes & Statistical Analyses

The primary outcomes were prevalence of SARS-CoV-2 infection by PCR and seroprevalence by laboratory-based antibody testing. Since we sampled the majority of the population of Bolinas (i.e., we obtained a sample without replacement from a finite population) we assumed data followed a hypergeometric distribution.

To further interpret the PCR results, we calculated the probability of observing x PCR positive cases, conditional on there being K true cases in the population of size N (of which we tested n).[20,21] Models also accounted for the sensitivity and specificity of the PCR test; we used values of 80% and 100%, respectively.[22,23]

We used a Bayesian modeling approach to estimate population seroprevalence based on the results of the two antibody testing platforms, accounting for test performance characteristics.[24] For validation data, we used the package insert data for the Abbott test[17] (1066 of 1070 negative controls tested negative; 88 of 88 PCR+ positive controls tested positive), and in-house validation of the ELISA test (95 of 95 negative controls tested negative; 42 of 44 positive controls tested positive). We note that test sensitivities were based on different patient populations (Abbott test based on hospitalized patients, ELISA based predominantly on ambulatory patients) and therefore not directly comparable. We first estimated seroprevalences independently by assay, and then estimated a single seroprevalence in a multinomial model which assumed the two assays to be conditionally independent.[25] For both scenarios we calculated positive predictive values based on estimated seroprevalence and test performance. All analysis was conducted using the R statistical software (http://cran.rproject.org) and the Stan programming language (http://mc-stan.org/). See Appendix 1 for more detailed explanation of statistical methods; additionally, code to reproduce all analyses are available at: https://github.com/EPPIcenter/bolinas-analysis.

### Ethics Statement

The study protocol was submitted to UCSF’s Institutional Review Board, and the study was deemed public health surveillance not requiring IRB oversight [IRB number 20–30636].

## RESULTS

Of 1,891 participants tested in Bolinas between April 20^th^ and April 24^th^, 1,312 were confirmed Bolinas residents, 76 were non-Bolinas resident first responders, essential workers, and their families, 47 were non-resident volunteers, and 456 were registered onsite (a mix of Bolinas residents, non-Bolinas first responders/essential workers). Based on aforementioned 2010 Census and 2018 ACS Bolinas population estimates, we calculated community ascertainment of greater than 80%. Demographic, epidemiologic, and symptom-related characteristics of survey respondents are listed in Table 1. Most participants were adults aged 18 and over (90%), with over one third aged 60 and older (35%). The majority of participants identified as White/Caucasian, with almost a third (31%) reporting annual household income less than $50,000. The vast majority of survey respondents reported wearing a mask (93%), and most estimated that they left their homes only 0–1 times weekly for work, food, or other reasons (see Table 1). Finally, while only 2% of participants had symptoms consistent with COVID-19 (e.g. fever, cough, muscle aches, severe fatigue, difficulty breathing, diarrhea, loss of smell and/or taste) on the day of testing, 31% reported having had at least one of the aforementioned symptoms in the month prior to testing.

**Table 1:**
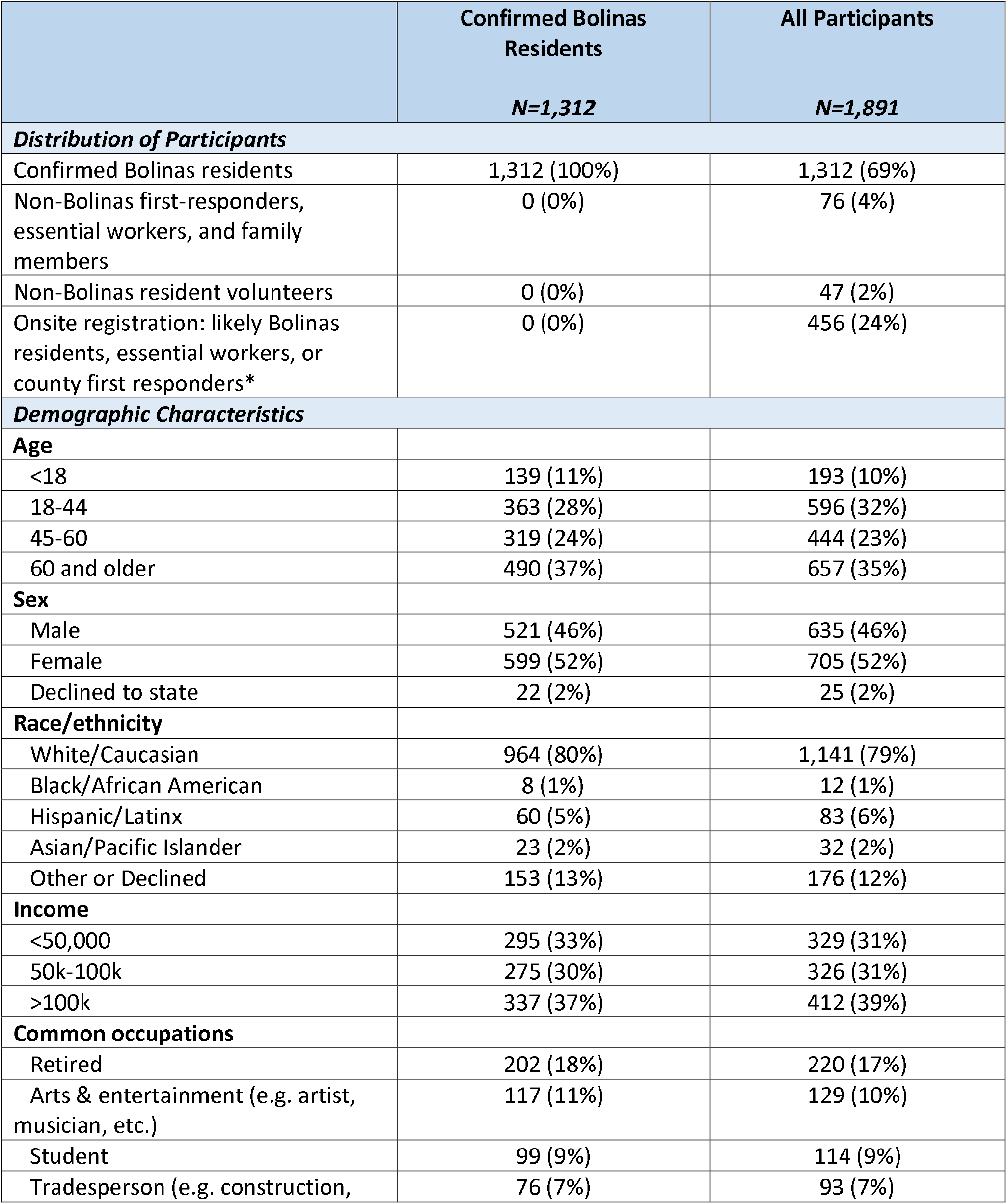
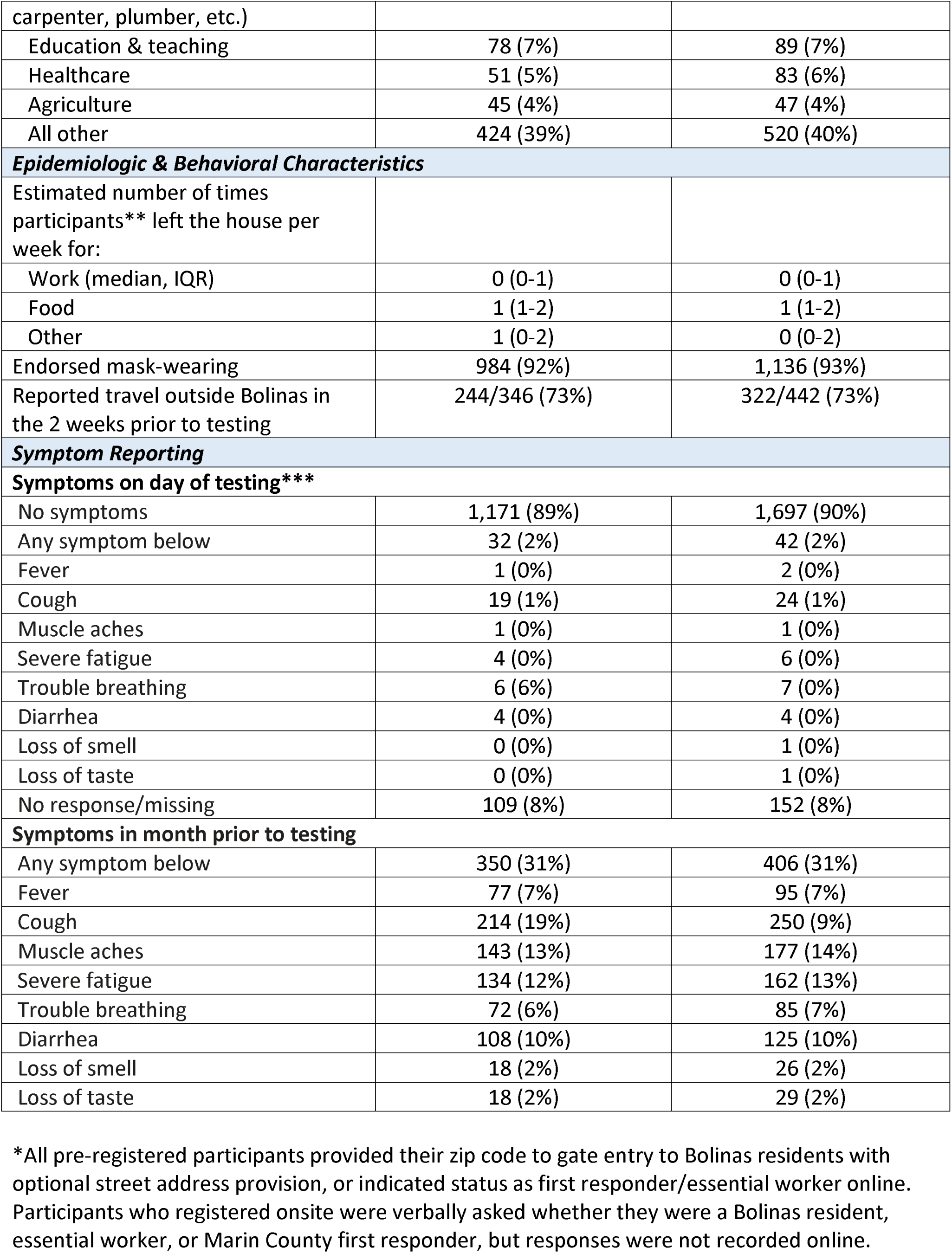
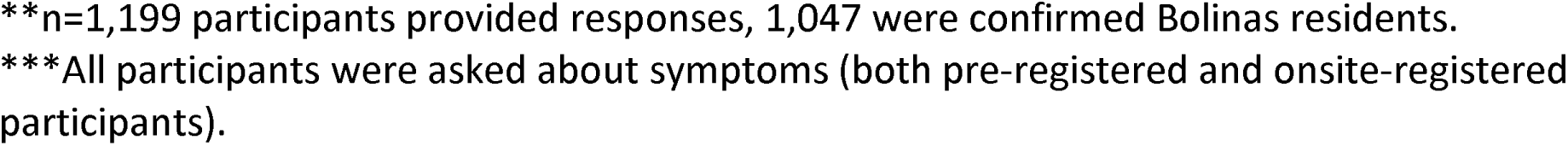
Demographic, Epidemiologic, and Symptom-related Characteristics of Participants

|  | Confirmed Bolinas Residents | All Participants |
| --- | --- | --- |
|  | *N=1,312* | *N=1,891* |
| ***Distribution of Participants*** |  |  |
| Confirmed Bolinas residents | 1,312 (100%) | 1,312 (69%) |
| Non-Bolinas first-responders, essential workers, and family members | 0 (0%) | 76 (4%) |
| Non-Bolinas resident volunteers | 0 (0%) | 47 (2%) |
| Onsite registration: likely Bolinas residents, essential workers, or county first responders* | 0 (0%) | 456 (24%) |
| ***Demographic Characteristics*** |  |  |
| **Age** |  |  |
| <18 | 139 (11%) | 193 (10%) |
| 18-44 | 363 (28%) | 596 (32%) |
| 45-60 | 319 (24%) | 444 (23%) |
| 60 and older | 490 (37%) | 657 (35%) |
| **Sex** |  |  |
| Male | 521 (46%) | 635 (46%) |
| Female | 599 (52%) | 705 (52%) |
| Declined to state | 22 (2%) | 25 (2%) |
| **Race/ethnicity** |  |  |
| White/Caucasian | 964 (80%) | 1,141 (79%) |
| Black/African American | 8 (1%) | 12 (1%) |
| Hispanic/Latinx | 60 (5%) | 83 (6%) |
| Asian/Pacific Islander | 23 (2%) | 32 (2%) |
| Other or Declined | 153 (13%) | 176 (12%) |
| **Income** |  |  |
| <50,000 | 295 (33%) | 329 (31%) |
| 50k-100k | 275 (30%) | 326 (31%) |
| >100k | 337 (37%) | 412 (39%) |
| **Common occupations** |  |  |
| Retired | 202 (18%) | 220 (17%) |
| Arts & entertainment (e.g. artist, musician, etc.) | 117 (11%) | 129 (10%) |
| Student | 99 (9%) | 114 (9%) |
| Tradesperson (e.g. construction, | 76 (7%) | 93 (7%) |
| carpenter, plumber, etc.) |  |  |
| Education & teaching | 78 (7%) | 89 (7%) |
| Healthcare | 51 (5%) | 83 (6%) |
| Agriculture | 45 (4%) | 47 (4%) |
| All other | 424 (39%) | 520 (40%) |
| ***Epidemiologic & Behavioral Characteristics*** |  |  |
| Estimated number of times participants** left the house per week for: |  |  |
| Work (median, IQR) | 0 (0-1) | 0 (0-1) |
| Food | 1 (1-2) | 1 (1-2) |
| Other | 1 (0-2) | 0 (0-2) |
| Endorsed mask-wearing | 984 (92%) | 1,136 (93%) |
| Reported travel outside Bolinas in the 2 weeks prior to testing | 244/346 (73%) | 322/442 (73%) |
| ***Symptom Reporting*** |  |  |
| **Symptoms on day of testing\*\*\*** |  |  |
| No symptoms | 1,171 (89%) | 1,697 (90%) |
| Any symptom below | 32 (2%) | 42 (2%) |
| Fever | 1 (0%) | 2 (0%) |
| Cough | 19 (1%) | 24 (1%) |
| Muscle aches | 1 (0%) | 1 (0%) |
| Severe fatigue | 4 (0%) | 6 (0%) |
| Trouble breathing | 6 (6%) | 7 (0%) |
| Diarrhea | 4 (0%) | 4 (0%) |
| Loss of smell | 0 (0%) | 1 (0%) |
| Loss of taste | 0 (0%) | 1 (0%) |
| No response/missing | 109 (8%) | 152 (8%) |
| **Symptoms in month prior to testing** |  |  |
| Any symptom below | 350 (31%) | 406 (31%) |
| Fever | 77 (7%) | 95 (7%) |
| Cough | 214 (19%) | 250 (9%) |
| Muscle aches | 143 (13%) | 177 (14%) |
| Severe fatigue | 134 (12%) | 162 (13%) |
| Trouble breathing | 72 (6%) | 85 (7%) |
| Diarrhea | 108 (10%) | 125 (10%) |
| Loss of smell | 18 (2%) | 26 (2%) |
| Loss of taste | 18 (2%) | 29 (2%) |

\*All pre-registered participants provided their zip code to gate entry to Bolinas residents with optional street address provision, or indicated status as first responder/essential worker online. Participants who registered onsite were verbally asked whether they were a Bolinas resident, essential worker, or Marin County first responder, but responses were not recorded online.
\*\*n=1,199 participants provided responses, 1,047 were confirmed Bolinas residents.
\*\*\*All participants were asked about symptoms (both pre-registered and onsite-registered participants).

### RT-PCR Results

Of 1,847 RT-PCR tests performed for active SARS-CoV-2 infection, 0 were positive. Using an estimated test sensitivity of 80%, conservatively assuming 80% of the community was sampled, and only including confirmed Bolinas residents, this corresponds to a population prevalence of 0.00048 (95% credible interval [CrI] 0.00001 – 0.00176). The calculated probability of observing 0 infections if there were truly 3 active infections in the community was < 5%, with probabilities dropping further for higher numbers of infections (Figure 1). Thus, it is likely that few if any active infections were present in the community at the time of sampling.

**Figure 1.**
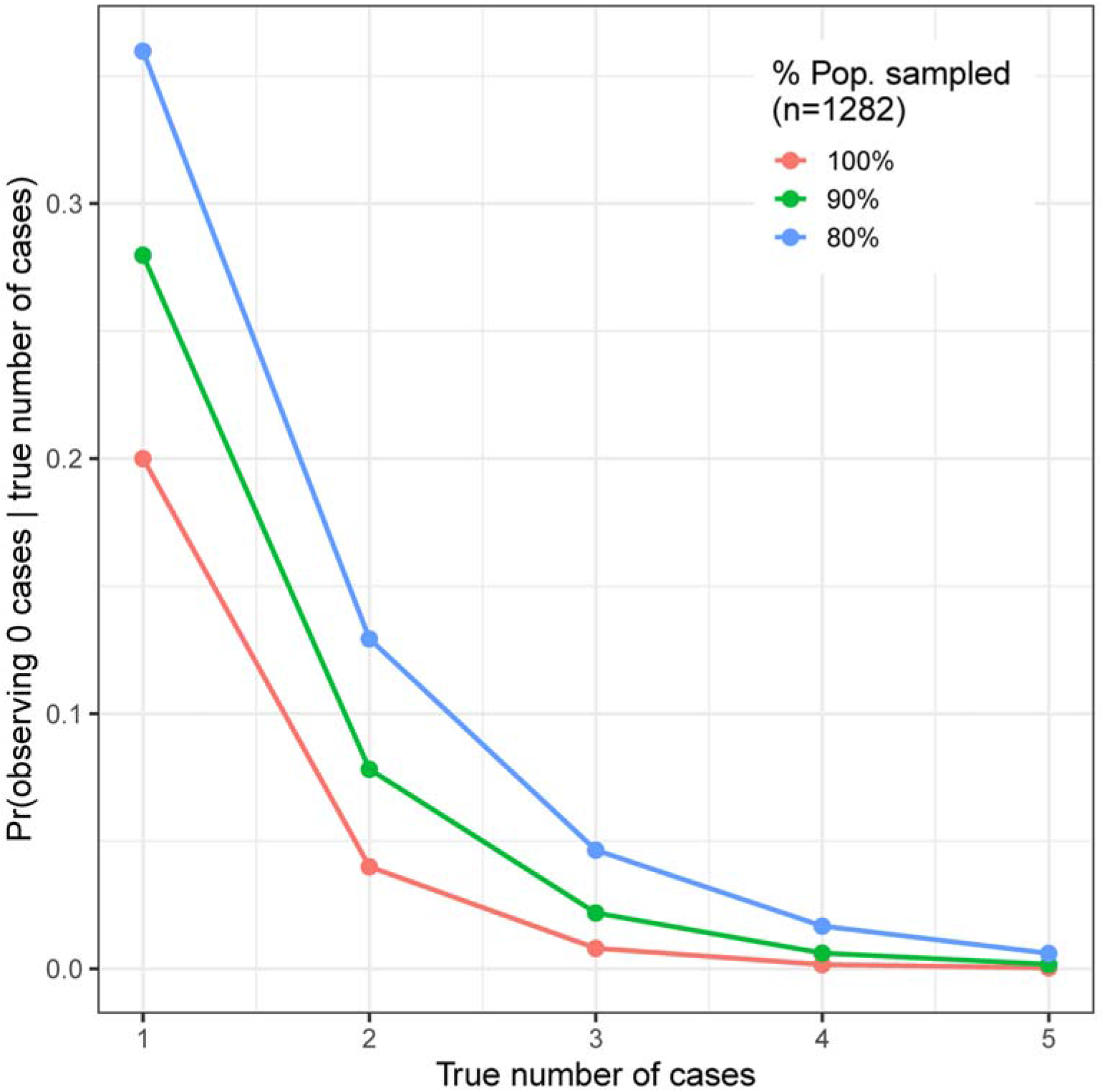
Probability of observing 0 cases given the true number of cases (y-axis), across a range of true numbers of cases (x-axis) and the proportion of the total population that was sampled (red, green, and blue lines). For example: assuming that we had sampled the entire population (red line), the probability of observing 0 cases if there truly had been 1 case is 0.2, or 20%; the probability of observing 0 cases if there truly had been 3 cases is 0.008.

### Antibody Results

Of 1,880 participants with antibody tests performed for prior SARS-CoV-2 infection, 12 participants had a positive result, and 8 of 12 with positive antibody results were confirmed Bolinas residents. Only one participant, a Bolinas resident, had a positive result on both antibody assays, while 8 others had a positive result on the ARCHITECT Abbott IgG test alone, and 3 other participants had a positive result on the ELISA IgG test alone (see Figure 2). ELISA titer was the highest for the sample also testing positive via the Abbott test, at > 1:100.

**Figure 2.**
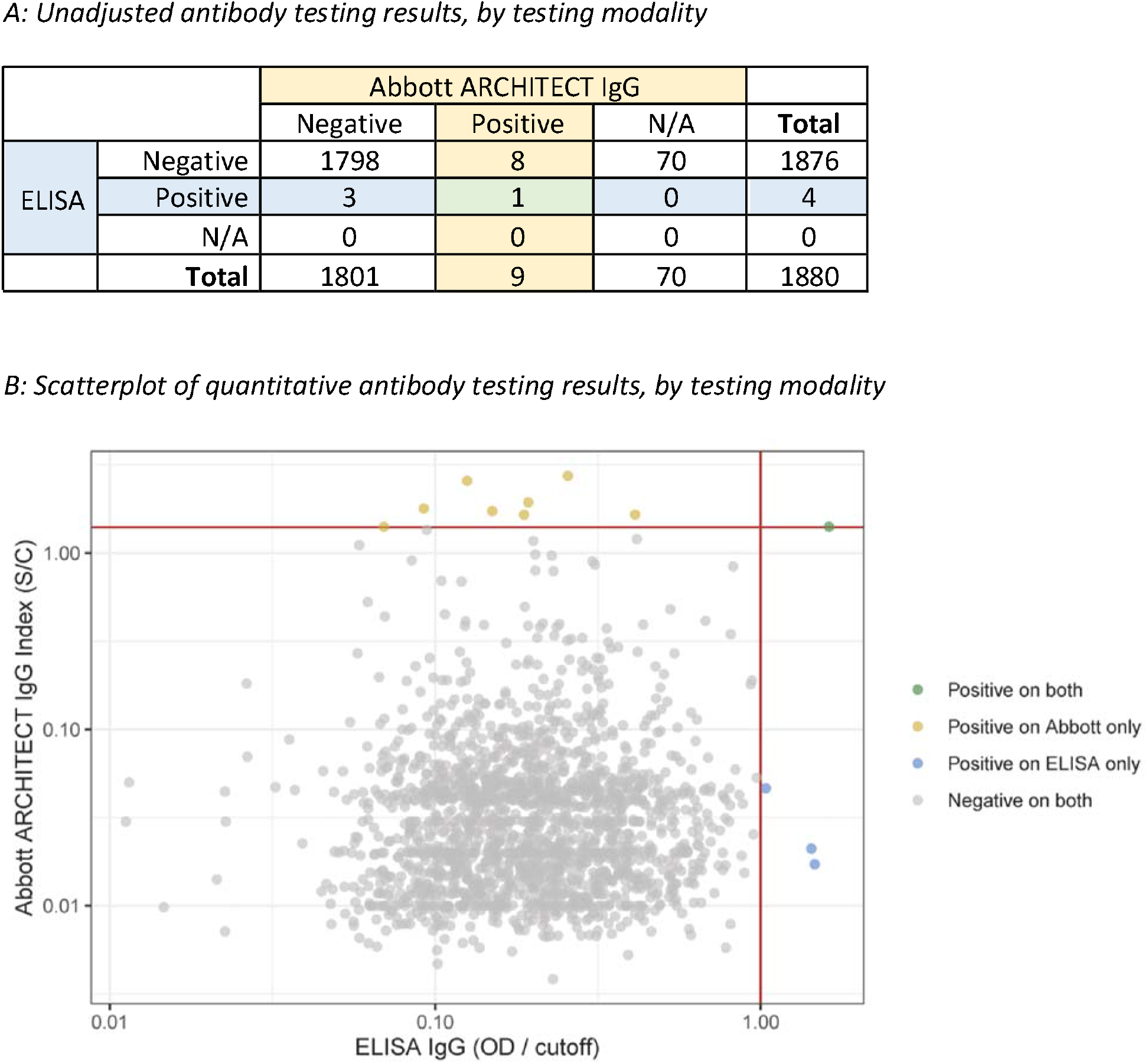
Panel A illustrates the number of specimens that were positive and negative, by testing modality. N/A indicates samples not run on Abbott assay due to insufficient plasma volume. Panel B illustrates the quantitative results of antibody tests that were run on both assays (n = 1,810) with colors denoting positive results. Abbott ARCHITECT IgG signal to cutoff values are shown on the Y-axis with ELISA IgG optical density to cutoff values on the X-axis.

Considering each test independently, correcting for the estimated sensitivity and specificities of the assays, and including only confirmed Bolinas residents, we estimated seroprevalence to be 0.29% (95% credible interval (CrI): 0.01%, 0.78%) by the Abbott assay and 0.23% (95% CrI: 0.01%, 0.62%) by the ELISA (Table 2). The wide range of the 95% CrI for these prevalence estimates reflects the high uncertainty in interpreting a positive test result in the setting of very low prevalence. Using the data from both tests, we estimated seroprevalence to be 0.16% (95% CrI: 0.02%, 0.46%). Importantly, the availability of two antibody results per sample sharply increased the positive predictive value (PPV): the probability of an individual testing positive on both Abbott and ELISA being truly infected was > 99%, whereas the PPV for just one of the two tests was low (< 2% for both configurations). If only one of the antibody tests were used, the PPV of a positive test would have been much lower and much more imprecise: between 44 – 63% with credible intervals that ranged from ∼3% to ∼98% (see Table 2).

**Table 2:**
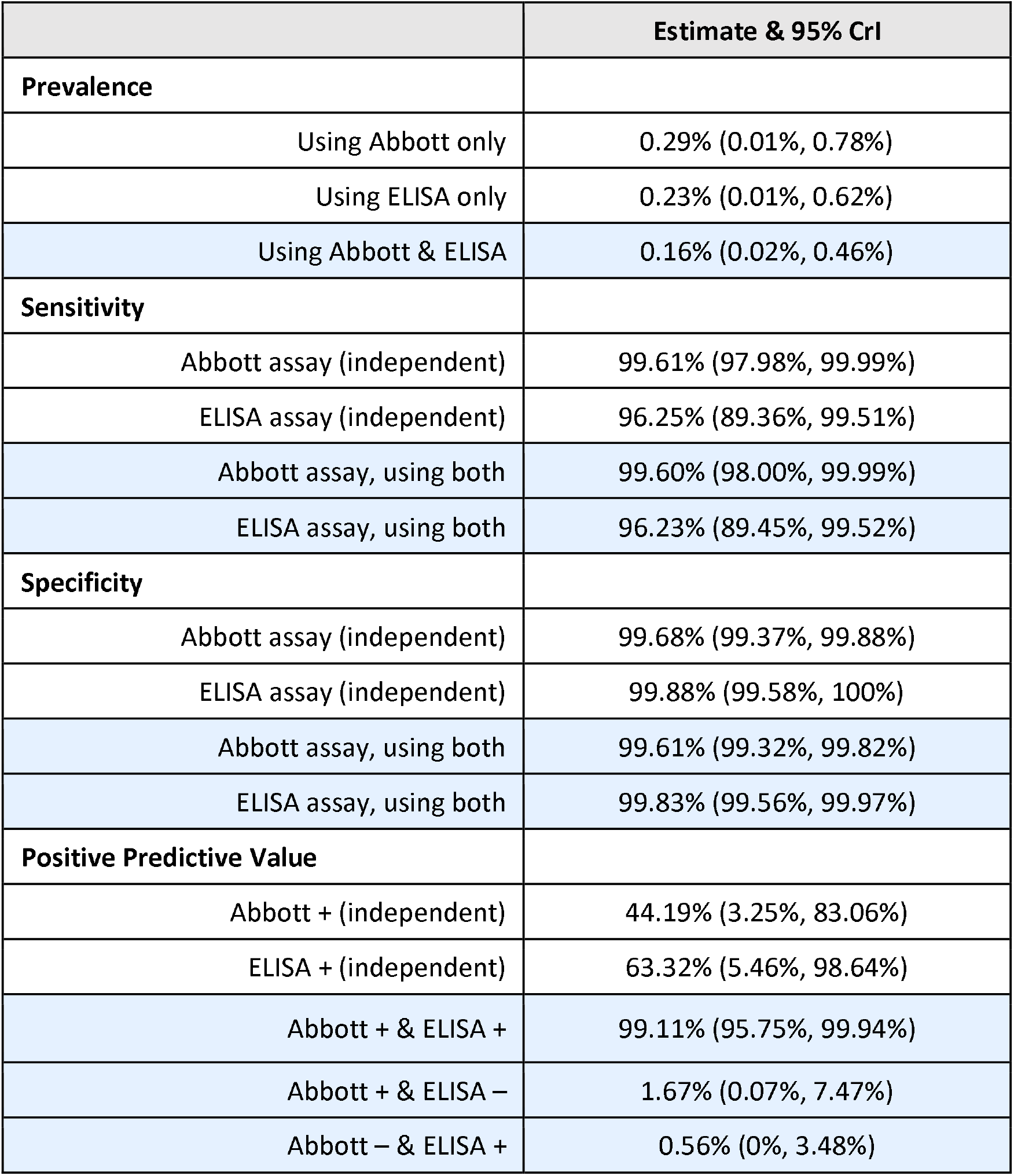
Modeled Prevalence, Sensitivity, Specificity and Positive Predictive Value based on Independent and Conjoined Antibody Testing Results

|  | Estimate & 95% CrI |
| --- | --- |
| **Prevalence** |  |
| Using Abbott only | 0.29% (0.01%, 0.78%) |
| Using ELISA only | 0.23% (0.01%, 0.62%) |
| Using Abbott & ELISA | 0.16% (0.02%, 0.46%) |
| **Sensitivity** |  |
| Abbott assay (independent) | 99.61% (97.98%, 99.99%) |
| ELISA assay (independent) | 96.25% (89.36%, 99.51%) |
| Abbott assay, using both | 99.60% (98.00%, 99.99%) |
| ELISA assay, using both | 96.23% (89.45%, 99.52%) |
| **Specificity** |  |
| Abbott assay (independent) | 99.68% (99.37%, 99.88%) |
| ELISA assay (independent) | 99.88% (99.58%, 100%) |
| Abbott assay, using both | 99.61% (99.32%, 99.82%) |
| ELISA assay, using both | 99.83% (99.56%, 99.97%) |
| **Positive Predictive Value** |  |
| Abbott + (independent) | 44.19% (3.25%, 83.06%) |
| ELISA + (independent) | 63.32% (5.46%, 98.64%) |
| Abbott + & ELISA + | 99.11% (95.75%, 99.94%) |
| Abbott + & ELISA – | 1.67% (0.07%, 7.47%) |
| Abbott – & ELISA + | 0.56% (0%, 3.48%) |

## DISCUSSION

Four weeks after shelter-in-place orders in a rural town comprised of mostly older adults with varied socioeconomic status, we found that active and prior SARS-CoV-2 infections were extremely rare, despite relative geographic proximity to urban areas with higher transmission. In this low transmission environment, use of two highly specific, independent antibody tests allowed for precise estimation of seroprevalence.

As the COVID-19 pandemic has continued, two things have become increasingly clear: 1) SARS-CoV-2 testing regardless of symptoms has proved very important, and 2) the penetration of SARS-CoV-2 has been alarmingly uneven in different populations. Universal testing in homeless shelters[26], prisons[27], nursing facilities[28], and hospitals[29] has demonstrated high rates of infection without concurrent symptoms. Similarly, in the limited existing community-based surveillance data published at time of this report, prior SARS-CoV-2 infection in an urban area of San Francisco was more common than passive case detection would have suggested.[30] In contrast, we found that SARS-CoV-2 infection was extremely uncommon, and potentially nonexistent, in our study setting. In light of rapidly expanding evidence supporting the effectiveness of mask-wearing and physical distancing[31–33], our findings of nearly zero infections in a community with high uptake of mask-wearing and adherence to shelter-in-place health orders support the effectiveness of these common public health measures. The high rate of turnout for testing is also indicative of notable community awareness and engagement that may have also facilitated consistent adherence. Other possible contributing factors include the Bay Area’s relatively few number of cases (at the time of testing) and the low population density in Bolinas. However, population density alone is not a sufficient explanation; while less publicized than urban epidemics, the devastating experience of Navajo Nation[34], and many other rural, native communities, stands in stark contrast to our data, illustrating that some rural communities may be particularly vulnerable to severe consequences of SARS-CoV-2 spread. Similarly in New York City, it was not the most population dense borough (Manhattan) that saw the highest number of cases, but rather the borough with the highest proportion of people of color who bore a disproportionate burden of poverty and other consequences of systemic racism that had the highest number of cases.[9] In summary, these data from Bolinas suggest that while rural communities may be vulnerable to severe consequences of SARS-CoV-2 given older age, high rates of poverty, and limited access to testing and medical care, this particular community was able to suppress the risk of infection four weeks into sheltering-in-place. Despite a rate of poverty higher than the national average, this community that sought to test itself with the self-reported goal of protecting its elders, whose constituents were also mostly able to shelter-in-place in a rural coastal setting and reported high uptake of mask-wearing, was able to remain close to free from infection.

Given the very low hypothesized prevalence of SARS-CoV-2 infection in this setting and to minimize the number of false positive results, we chose to test each sample using two specific, laboratory-based tests evaluating responses to different viral proteins of the virus. The implementation and interpretation of SARS-CoV-2 antibody testing has been complicated for a number of reasons including: different indications for antibody testing (e.g., diagnosing active infection, confirming convalescent infection for plasma donation, and seroprevalence) that prioritize different optimal test characteristics, use of laboratory and non-laboratory based methods with wide variation in accuracy, and sparse and poorly characterized validation data, among other concerns.[12,35] High specificity of an assay is particularly important when performing serosurveillance in areas with lower transmission, since false positives will be a higher proportion of the total positives. The estimated specificity of our two independent tests were both > 99.5%, but given the extremely low prevalence of infection in this community, the positive predictive value of either test alone remained low. By jointly analyzing the results of these two tests, we were able to more precisely estimate seroprevalence and obtain a high positive predictive value when both tests were positive. In this study, given only one individual was likely to have been truly exposed, we were not able to use these data to systematically risk factors for infection. However, having a high positive predictive value may allow better estimation of risk factors in other studies and may allow for more meaningful communication of results to individuals. Given aforementioned issues regarding interpretation of antibody data in low prevalence areas[36], using multiple orthogonal tests or at least multiple antigenic targets may be a useful strategy to improve the overall performance and augment the utility of serosurveillance moving forward.

Our study was subject to important limitations. For nearly all SARS-CoV-2 antibody tests, the true sensitivity when applied to a community-based sample where the majority of infected individuals will have experienced mild or asymptomatic SARS-CoV-2 infections is unknown, as test performance characteristics have generally been calculated based on severe infections only.[37–39] We note that here, at least, sensitivity for the ELISA was evaluated largely on mild (though not asymptomatic) infections. In this particular population with low prevalence of active or prior infection, false negative results were less of a concern and would have minimally changed seroprevalence estimates. Next, it is possible that we sampled a biased subset of the community, e.g. with certain demographic groups or those experiencing illness less likely to leave their homes for testing. However, we estimated relatively high community ascertainment (>80%) and also tested home-bound participants, mitigating this factor. Finally, not all participants completed the epidemiologic survey, which may have introduced selection bias in that those who elected to share information about mask-wearing and movement during shelter-in-place were more likely to report socially desirable values. However, community members reported anecdotal observations consistent with our overall study findings.

In conclusion, active and prior SARS-CoV-2 infections were rare in this rural town with a high uptake of mask-wearing and compliance with shelter in place directives, despite relative proximity to urban areas with significantly higher transmission. Use of two independent, highly accurate antibody tests methods allowed for a more precise estimate of seroprevalence and higher positive predictive value than either test alone.

## Data Availability

See Appendix 1 for more detailed explanation of statistical methods; additionally, code to reproduce all analyses are available at: https://github.com/EPPIcenter/bolinas-analysis.

## Acknowledgements

We would like to acknowledge the significant contribution to this work made by the following persons: 1) Dr. Matt Willis of the Marin County Public Health Department for his support of this project within his jurisdiction, 2) Jacqueline Martinez and Andrew Kobylinski for their considerable technical expertise in operationalizing multiple aspects of this project, 3) Ana Vallari, Barb Harris, Ana Olivo, and Chris Lark at Abbott Laboratories for their contribution to generating serology data.

## 1 Appendix

### SUPPLEMENTARY METHODS

#### 1A. Testing the upper bound on the true number of infections

To test the upper bound on the true number of infections (via PCR), we calculated the probability of observing *x* = 0 cases, conditional on there being exactly *K* cases in our population of size *N* (of which we tested *n*). We assume that the sensitivity (*Se*) and specificity (*Sp*) of the diagnostic test are known and fixed at 80% and 100%, respectively. We assume that the n individuals were sampled at random and so are representative of the entire population N. We use the hypergeometric distribution since we have a finite population size so trials are not independent, and binomial distributions to account for *Se* and *Sp*.

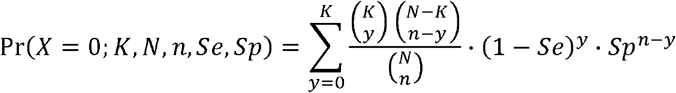

Code available at: https://github.com/EPPIcenter/bolinas-analysis/blob/master/1A_Figure_1.R.

#### 1B. Estimation of PCR prevalence

We estimated PCR prevalence in two ways: first, by using the binomial distribution to model prevalence (p) and second, by using the hypergeometric distribution to model prevalence. The latter is more appropriate in this scenario since testing was performed on a large proportion of the total population (upwards of 80%), so the hypergeometric distribution will yield narrower estimates of uncertainty. We again assume that the Se and Sp of the diagnostic test are known and fixed at 80% and 100%, respectively. As the binomial distribution is more frequently used for prevalence estimation, we provide Stan code to implement both models.

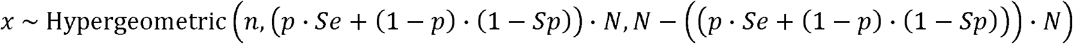

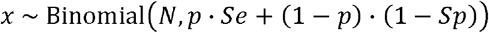

Code available at: https://github.com/EPPIcenter/bolinas-analysis/blob/master/1B.1_PCR_prevalence_HGM.R https://github.com/EPPIcenter/bolinas-analysis/blob/master/1B.2_PCR_prevalence_binomial.R

#### 2A. Estimation of seroprevalence (one test)

We used the binomial distribution to estimate seroprevalence (p) separately for each assay j. We use the binomial distribution, which will yield conservative intervals as compared to the hypergeometric distribution used as above. The difference from the estimation of PCR prevalence is that we also estimate Se and Sp of the assays using validation data. The positive predictive values (PPV) are calculated directly from the estimates.

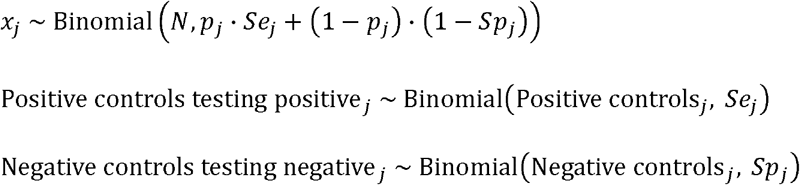

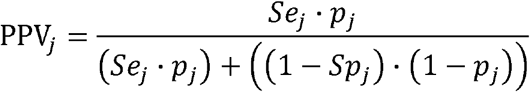

Code available at: https://github.com/EPPIcenter/bolinas-analysis/blob/master/2A_Ab_prevalence_separate_binomial.R

#### 2B. Estimation of seroprevalence (two tests)

We extended the approach in 2A above to jointly model the results of both assays, using the multinomial distribution as the generalization of the binomial distribution and now estimating a single seroprevalence p. We necessarily assumed conditional independence between the two assays j and k. For samples that were only tested on one platform, we allowed them to contribute to the binomial likelihood for the platform that they were tested on. The positive predictive values, which are now based on the results of both assays, are calculated directly from the estimates.

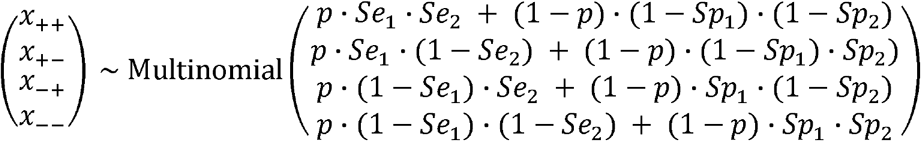

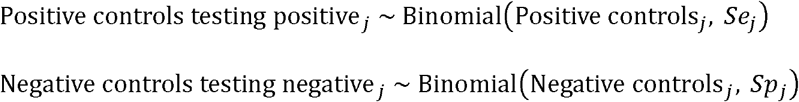

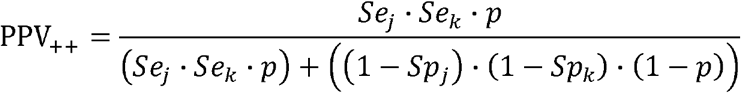

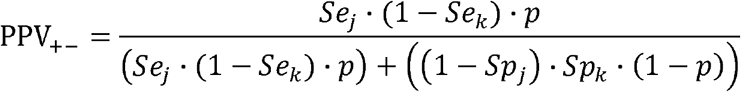

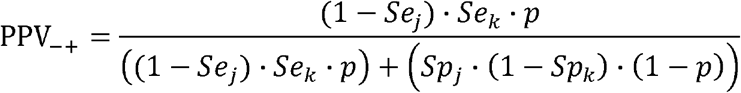

Code available at: https://github.com/EPPIcenter/bolinas-analysis/blob/master/2B_Ab_prevalence_joint_multinomial.R

